# An Interpretable DSP Architecture for ECG Rhythm and Premature Ventricular Contraction Detection in MATLAB/Simulink

**DOI:** 10.64898/2026.11.16.26368189

**Authors:** Abdullah Atif, Mst. Jannatul Ferdous

**Affiliations:** Department of Computer Science and Engineering, Varendra University, Rajshahi, Bangladesh

**Keywords:** Electrocardiogram, Premature ventricular contraction, QRS detection, Digital signal processing, MATLAB/Simulink, MIT-BIH Arrhythmia Database

## Abstract

Electrocardiogram (ECG) monitoring requires reliable signal preprocessing and interpretable detection of abnormal cardiac activity while maintaining low algorithmic complexity. This study presents an integrated digital signal processing architecture developed in MATLAB/Simulink for ECG preprocessing, QRS and R-peak detection, heart-rate estimation, rhythm classification, and premature ventricular contraction (PVC) detection. Real ECG recordings from the MIT-BIH Arrhythmia Database were used for development and evaluation. The preprocessing stage employs minimum-order elliptic IIR high-pass and low-pass filtering, followed by differentiation, squaring, moving-window integration, thresholding, and R-peak detection. RR intervals derived from detected R peaks are used for instantaneous heart-rate calculation and classification of normal rhythm, bradycardia, and tachycardia. PVC detection uses two interpretable beat-level features, an RR-interval ratio and a QRS-energy ratio, combined through fixed rule-based logic. Three initial 10-s windows containing 18 annotated PVCs were used during algorithm development and threshold selection. After the detector parameters were frozen, evaluation was performed on nine additional non-overlapping 10-s windows containing 56 expert-annotated PVCs from MIT-BIH Records 106, 119, and 200. Using a ±100 ms event-matching tolerance, the held-out evaluation produced 40 true positives, 9 false positives, and 16 false negatives, corresponding to 71.43% sensitivity, 81.63% precision, and 76.19% F1 score. Across the same nine windows, the R-peak detector achieved 94.39% sensitivity, 99.02% positive predictivity, 96.65% F1 score, and approximately 16.91 ms mean absolute timing error. The results demonstrate the feasibility and limitations of a lightweight and explainable streaming ECG-processing architecture and motivate future work incorporating adaptive decision thresholds and broader patient-independent validation.

## 1. Introduction

Cardiovascular monitoring frequently relies on the electrocardiogram (ECG), which provides a non-invasive representation of the electrical activity of the heart. Automated ECG processing is particularly important for ambulatory and long-duration monitoring because manual inspection of extended recordings is time-consuming. Among the abnormalities encountered in ECG recordings, premature ventricular contractions (PVCs) are ectopic ventricular beats whose automated identification has received considerable research attention. PVC detection has therefore been investigated using approaches ranging from relatively simple timing-based algorithms to model-based and contemporary deep-learning systems [5]–[8].

Reliable detection of abnormal beats depends first on robust identification of the QRS complex and the associated R peaks. Pan and Tompkins established a real-time signal-processing framework based on band-limited filtering, differentiation, squaring, moving-window integration, and decision logic. The method reported 99.3% QRS detection on the MIT-BIH Arrhythmia Database [1]. These principles remain attractive for systems in which computational transparency and low-latency implementation are important.

PVC classification can exploit temporal and morphological changes caused by abnormal ventricular activation. Cuesta et al. proposed a lightweight PVC detector based on RR-interval information, using prematurity and compensatory-pause characteristics followed by linear classification, and reported 90.13% sensitivity and 82.52% specificity [5]. More sophisticated alternatives have incorporated ECG morphology through model-based Bayesian estimation. Sayadi et al. proposed a wave-based Bayesian framework for robust PVC detection using dynamic ECG waveform tracking and an extended Kalman filter [6].

More recent methodsincreasingly employ learned representations. Deep metric learning combined with k-nearest-neighbor classification has been proposed to automatically extract discriminative PVC features and reduce dependence on manually designed features [7]. End-to-end deep neural networks have also been evaluated for PVC detection across multiple Holter databases [8]. Such methods can provide strong classification capability, but their learned representations and computational requirements differ substantially from classical DSP implementations designed for transparent block-level analysis.

MATLAB/Simulink provides a suitable environment for this type of system because ECG input, discrete-time filtering, QRS processing, decision logic, validation, and dashboard visualization can be integrated within a model-based signal-processing workflow. The objective of this study is therefore not to claim state-of-the-art PVC accuracy but to develop and evaluate an integrated, interpretable, and low-complexity ECG DSP architecture in MATLAB/Simulink.

The principal contributions are: (1) an integrated Simulink ECG-processing architecture; (2) a low-latency IIR preprocessing strategy adopted after FIR-delay analysis; (3) transparent RR-derived rhythm analysis for normal, bradycardic, and tachycardic states; (4) a lightweight PVC detector using RR-ratio and QRS-energy-ratio features; and (5) annotation-based development and held-out evaluation, in which three initial 10-s windows containing 18 PVCs were used during development and threshold selection, and nine additional non-overlapping 10-s windows containing 56 annotated PVCs were used for evaluation without record-specific retuning.

## 2. Related work

Automated ECG analysis has progressed from deterministic digital signal-processing algorithms to statistical learning and deep neural networks. Nevertheless, QRS localization remains fundamental because beat timing provides the basis for RR-interval analysis, heart-rate estimation, and beat-level arrhythmiacharacterization. Thereal-time QRS detector of Pan and Tompkinsremains a canonical low-complexity reference [1].

For PVC detection, temporal information contained in consecutive RR intervals provides a computationally inexpensive source of discriminatory information. Cuesta et al. demonstrated PVC detection from RR intervals using prematurity and compensatory-pause features [5]. However, reliance on RR information alone can be difficult when abnormal beats are not strongly premature or rhythm variation produces similar RR changes.

Morphological modeling provides another strategy. Sayadi et al. developed a Bayesian ECG framework using dynamic waveform modeling and extended Kalman filtering, enabling denoising, waveform tracking, and PVC detection [6]. Contemporary research increasingly employs learned representations; deep metric-learning methods and end-to-end deep neural networks can automatically derive discriminative features but introduce training, storage, optimization, and interpretability considerations [7], [8].

The present work occupies the low-complexity and interpretable end of this design spectrum. Rather than competing directly with high-dimensional learned classifiers, the proposed system combines deterministic ECG preprocessing, QRS/R-peak detection, RR-based rhythm analysis, and beat-level PVC classification using an RR ratio and QRS-energy ratio. All processing stages are integrated within a single Simulink model, allowing intermediate signals and decision states to remain observable.

## 3. Materials and methods

### 3.1. ECG dataset

Experiments were conducted using recordings from the MIT-BIH Arrhythmia Database distributed through PhysioNet. The database contains 48 approximately 30 -min two-channel ambulatory ECG recordings obtained from 47 subjects. Signals were digitized at 360 samples/s per channel, and approximately 110,000 beat annotations were produced through independent expert review and adjudication [2]–[4].

The modified limb lead II (MLII) signal was used as the primary processing channel whenever available because the proposed architecture was developed as a single-channel ECG monitoring system. Selected segments from Records 100, 101, and 213 were used to demonstrate normal, bradycardic, and tachycardic rhythm operation, respectively. PVC development and evaluation used segments from Records 106, 119, and 200.

Three initial 10-s windows, Record 106 at 88–98 s, Record 119 at 20–30 s, and Record 200 at 0–10 s, contained a total of 18 annotated PVCs and were used during algorithm development and threshold selection. These windows are therefore reported as development data rather than held-out evaluation data.

Following parameter selection, all active detector thresholds were frozen. After the development windows were excluded, each record was divided into consecutive, non-overlapping 10-s candidate windows, and the number of expert-annotated PVCs in each window was computed from the annotation file. Candidate windows were ranked in descending order of PVC count, and evaluation windows were selected from the highest-density candidates subject to a predefined computational limit on the total number of events and with the aim of including more than one record. Selection stopped when the planned PVC-event budget was reached. This targeted sampling procedure increased the number of ventricular events available for stress testing within the practical simulation budget. The resulting evaluation therefore intentionally over-represents PVC-rich intervals and should not be interpreted as prevalence-weighted whole-record performance.

The final held-out evaluation contained 56 annotated PVCs across nine 10 -s windows from Records 106, 119, and 200. Each window was processed independently using identical sampling, filtering, feature-extraction, and decision parameters. ECG segments and corresponding annotation signals were converted to MAT-files and supplied through Simulink From File blocks.

### 3.2. Overall system architecture

The architecture comprises ECG file acquisition, IIR preprocessing, QRS enhancement and R-peak detection, RR-interval and heart-rate calculation, heart-rate-based rhythm classification, PVC feature extraction, rule-based PVC detection, reference-annotation validation, and dashboard-based monitoring (Fig. 1). Following R-peak detection, the processing pipeline separates into rhythm-analysis and PVC-analysis branches.

**Fig 1.**
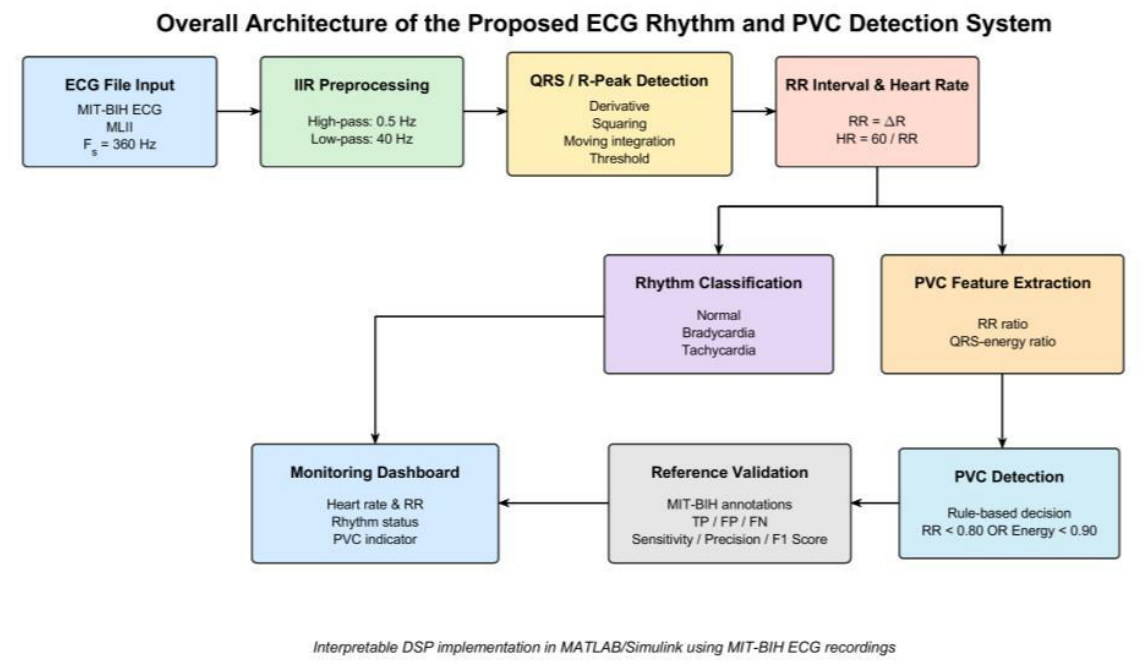
Overall system architecture of the proposed ECG rhythm and PVC detection system.

### 3.3. ECG preprocessing

The raw ECG contains low-frequency baseline variation and higher-frequency components that may interfere with reliable QRS detection. The preprocessing stage there fore employs cascaded digital high-pass and low-pass filters. During initial development, minimum-order FIR filters were evaluated. The high-pass design generated a filter length of 2613 coefficients (order 2612). For an approximately linear-phase FIR implementation, the corresponding group delay was:

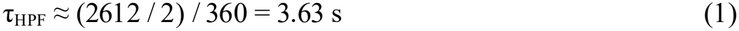

The low-pass FIR length was 246 and contributed approximately 0.34 s additional group delay. The combined delay, approaching 4 s, was incompatible with responsive monitoring.

Consequently, the final preprocessing architecture employed minimum-order elliptic IIR filters. Baseline-wander suppression used a high-pass filter implemented as three Direct-Form I second-order sections. At F_s_ = 360 Hz, the high-pass filter used a 0.1-Hz stopband edge, 0.5-Hz passband edge, 80-dB minimum stopband attenuation, and 0.1-dB maximum passband ripple. MATLAB reported the resulting filter as stable and non-linear-phase.

High-frequency suppression used a minimum-order elliptic low-pass filter implemented as six Direct-Form I second-order sections. The low-pass design used a 40-Hz passband edge, 45-Hz stopband edge, 80-dB minimum stopband attenuation, and 0.1-dB maximum passband ripple at the same 360-Hz sampling rate. The−3-dB frequency reported by the filter-analysis tool was approximately 40.385 Hz.

These IIR filters replaced the initially evaluated minimum-order FIR stages and were adopted as the final streaming-oriented preprocessing implementation. No claim of hardware real-time execution is made in this study. The effect of preprocessing in the time and frequency domains is shown in Figs. 2 and 3.

**Fig 2.**
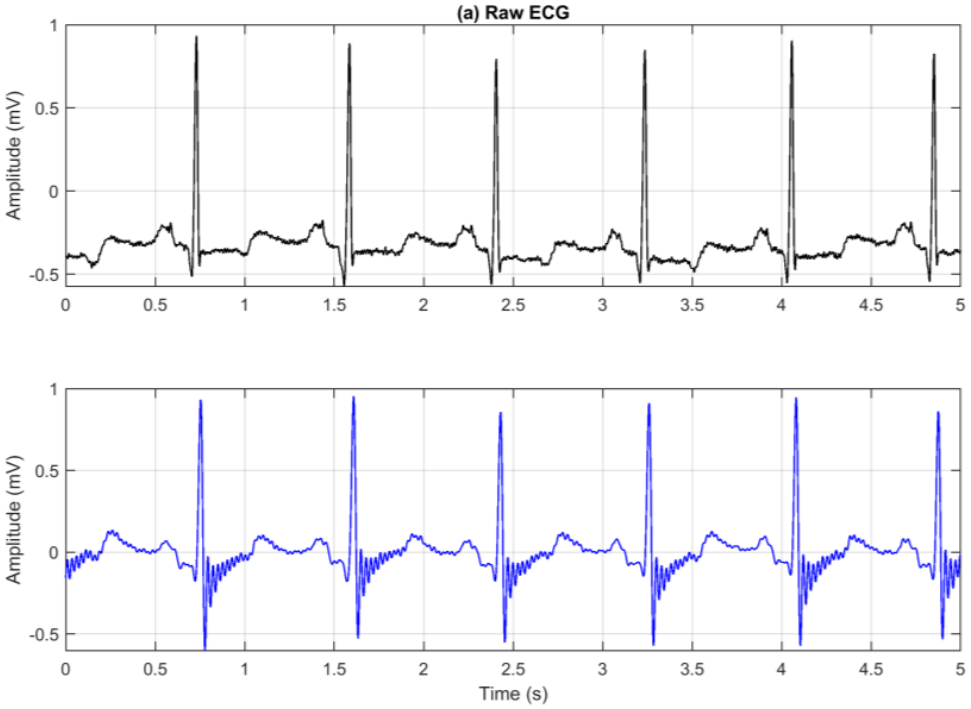
Raw (top) and preprocessed (bottom) MLII ECG segment from MIT-BIH Record 100.

**Fig 3.**
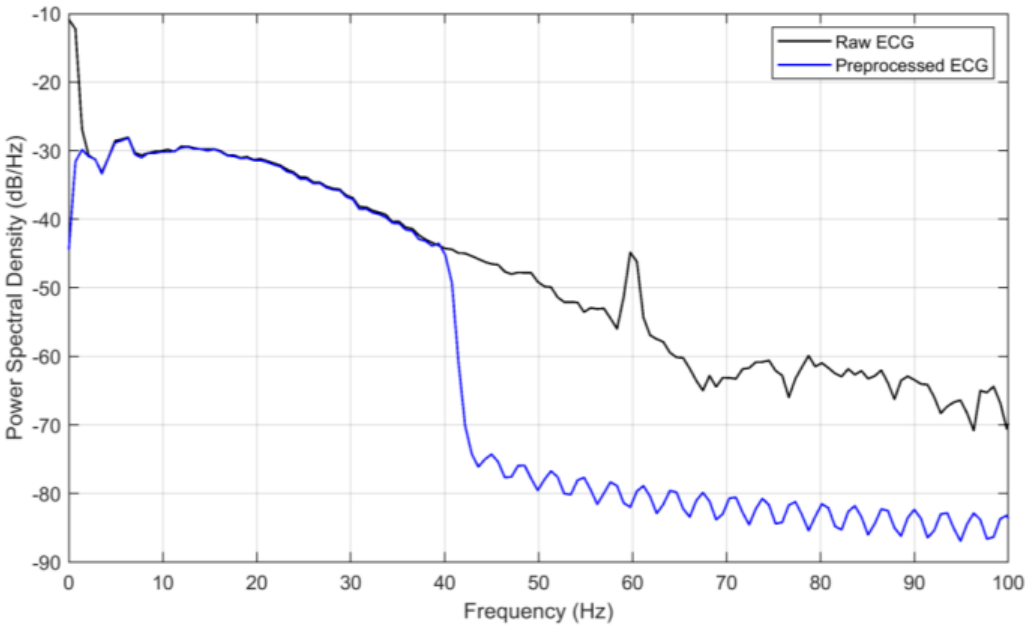
Power spectral density comparison of raw and preprocessed ECG using Welch estimation.

### 3.4. QRS and R-peak detection

The QRS detector follows a low-complexity DSP sequence inspired by the classical Pan–Tompkins method [1]. The preprocessed signal x[n] is first differentiated:

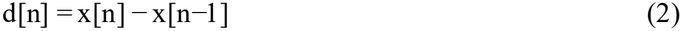

The derivative output is squared to emphasize high-slope QRS components:

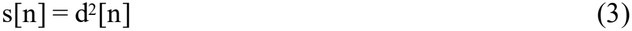

A 54-sample moving-window integration stage corresponds to approximately 150 ms at 360 Hz:

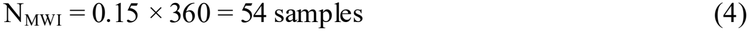

The integrated signal is compared against a decision threshold to identify QRS regions, and a rising-edge detector converts each threshold crossing into a discrete beat event. For publication visualization only, R-peak markers in Fig. 4 were localized to the nearest ECG maximum within ±100 ms; this did not alter the Simulink detector or quantitative evaluation.

**Fig 4.**
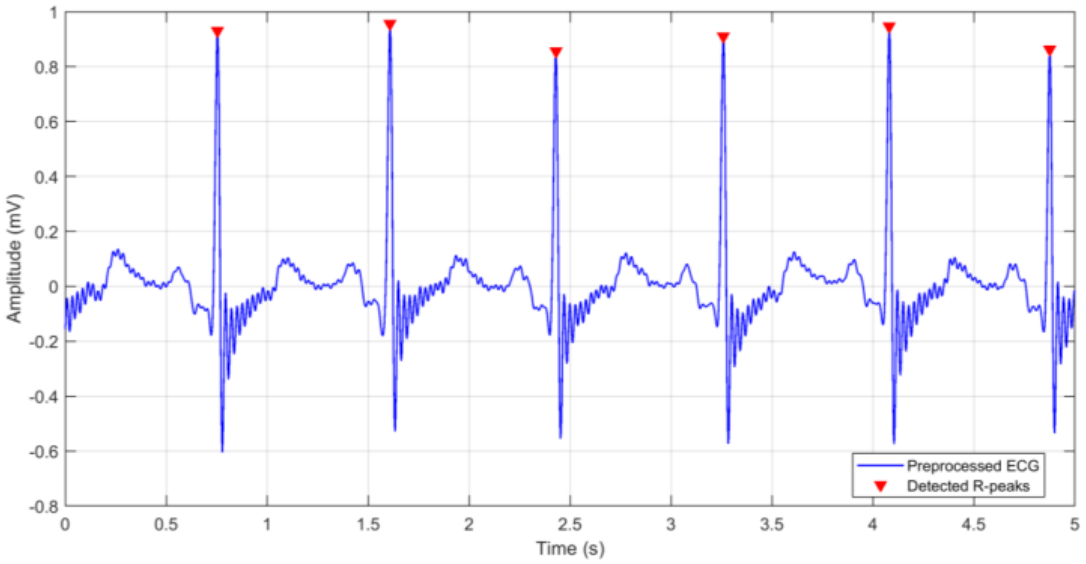
R-peak detection in a preprocessed MLII ECG segment from MIT-BIH Record 100.

The final integrated-QRS decision threshold was fixed at 0.005. All ECG processing used a fixed-step discrete sample period of 1/360 s. These settings were maintained unchanged during the held -out evaluation.

### 3.5. RR interval and heart-rate estimation

A free-running discrete sample counter operates at the ECG sampling frequency. At each detected R peak, a triggered subsystem records the current counter value. The interval between successive R peaks is:

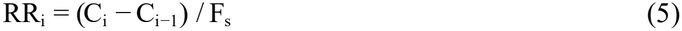

Instantaneous heart rate is calculated as:

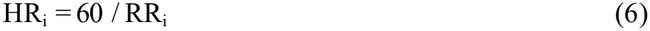

### 3.6. Rhythm classification

Three rhythm states were implemented using transparent heart-rate thresholds. Bradycardia was identified for HR < 60 beats/min, tachycardia for HR > 100 beats/min, and otherwise the signal was classified as normal rhythm. Representative validation examples used Records 100, 101, and 213, with measured values of approximately 69.9, 59.67, and 114.3 beats/min, respectively.

### 3.7. PVC feature extraction

#### 3.7.1. RR-interval ratio

For each detected beat, the current RR interval was compared with a recent RR baseline computed using a moving average of the previous eight RR intervals. The RR ratio was defined as:

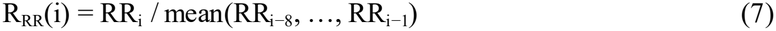

Here RR_i_ denotes the current RR interval. A beat satisfied the prematurity condition when R_RR_ < 0.80. The moving-average length and 0.80 threshold were selected during development and remained fixed during held-out evaluation.

#### 3.7.2. QRS-energy ratio

The morphology-related feature was derived from the squared derivative signal already available within the QRS-processing pathway. A 54-sample moving window generated an energy-related QRS measure, which was captured at each detected beat. The ratio between the current and previous captured values was defined as:

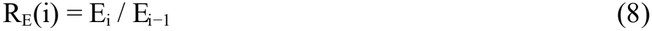

The final classifier retained the low-energy condition R_E_ < 0.90. This threshold was fixed before held-out evaluation.

### 3.8. PVC decision logic

The final PVC decision is a transparent OR rule:

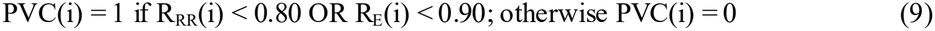

Alternative development branches included QRS-width estimation, high-energy detection, RR-change analysis, and compensatory-pause confirmation. These branches were excluded from the final active classifier because they did not consistently improve validation performance. Detector parameters were frozen before held-out evaluation.

### 3.9. Simulink implementation

The complete processing chain was implemented in MATLAB/Simulink R2019a using discrete signal-processing blocks. ECG and annotation segments were supplied using MAT-files through From File blocks. The model used a fixed-step discrete solver with a fundamental sample period of:

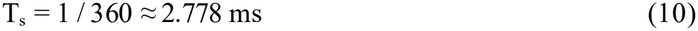

The architecture processes samples in streaming discrete-time form and includes a Simulink Dashboard for visualization of heart rate, RR interval, rhythm state, and PVC state. The present work does not report measured execution time, processor utilization, or hardware latency; consequently, the implementation is described as real-time-oriented or streaming-capable, rather than as experimentally verified real-time hardware operation.

### 3.10. Evaluation protocol and performance metrics

PVC performance was evaluated against expert annotations. A detected event was a true positive when it occurred within ±100 ms of an unmatched reference PVC; each reference event could be matched at most once. False positives were unmatched detections and false negatives were unmatched reference PVCs.

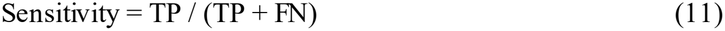

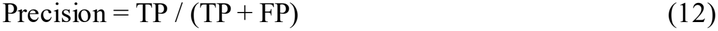

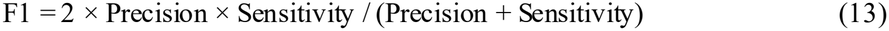

Development and final evaluation were explicitly separated. The three development windows contained 18 expert-annotated PVCs. After selection of the RR-ratio, QRS-energy, and QRS-detection thresholds, all parameters were frozen.

Held-out evaluation used nine additional non-overlapping 10-s windows containing 56 expert-annotated PVCs. These windows were selected deterministically by annotated PVC count to increase event density for stress testing. No thresholds were modified after the held-out evaluation began.

The upstream R-peak detector was evaluated separately on the same nine held-out windows using expert beat annotations and the same±100 ms matching tolerance. R-peak sensitivity, positive predictivity (TP / (TP + FP)), F1 score, and mean absolute timing error of matched detections were calculated independently of the PVC classifier; the pooled timing error was weighted by the number of true positives in each window.

## 4. Results

### 4.1. ECG preprocessing and spectral characteristics

The preprocessing stage reduced baseline variation while retaining visually distinct QRS complexes. Fig. 2 compares representative raw and preprocessed MLII ECG, whereas Fig. 3 shows Welch power spectral-density estimates. The preprocessed signal exhibits substantial attenuation beyond the approximately 40-Hz low-pass region. Replacing the original FIR stages with IIR implementations eliminated the several-second development latency and enabled beat detection throughout the windows.

### 4.2. R-peak and heart-rate analysis

The derivative, squaring, moving-window integration, and thresholding stages generated discrete R-peak events from the preprocessed ECG. A representative waveform is shown in Fig. 4.

Quantitative R-peak evaluation was conducted across the same nine held-out windows used for the final PVC analysis (Table 1). The evaluation contained 107 expert reference beats. The detector produced 101 true positives, 1 false positive, and 6 false negatives, corresponding to 94.39% sensitivity, 99.02% positive predictivity, and 96.65% F1 score. The true-positive-weighted mean absolute R-peak timing error was approximately 16.91 ms.

**Table 1.** R-peak detection performance in the nine held-out windows (±100 ms matching tolerance; MAE, mean absolute timing error).

| Record | Window (s) | Ref. beats | TP | FP | FN | Sens. (%) | PPV (%) | F1 (%) | MAE (ms) |
|---|---|---|---|---|---|---|---|---|---|
| 119 | 190–200 | 11 | 11 | 0 | 0 | 100.00 | 100.00 | 100.00 | 14.90 |
| 119 | 290–300 | 11 | 11 | 0 | 0 | 100.00 | 100.00 | 100.00 | 10.10 |
| 119 | 460–470 | 11 | 11 | 0 | 0 | 100.00 | 100.00 | 100.00 | 9.34 |
| 119 | 760–770 | 11 | 11 | 1 | 0 | 100.00 | 91.67 | 95.65 | 11.11 |
| 119 | 960–970 | 11 | 11 | 0 | 0 | 100.00 | 100.00 | 100.00 | 10.35 |
| 119 | 1130–1140 | 12 | 12 | 0 | 0 | 100.00 | 100.00 | 100.00 | 11.34 |
| 200 | 1100–1110 | 14 | 12 | 0 | 2 | 85.71 | 100.00 | 92.31 | 27.78 |
| 200 | 70–80 | 14 | 14 | 0 | 0 | 100.00 | 100.00 | 100.00 | 24.21 |
| 106 | 130–140 | 12 | 8 | 0 | 4 | 66.67 | 100.00 | 80.00 | 35.76 |
| **Pooled** | **9 windows** | **107** | **101** | **1** | **6** | **94.39** | **99.02** | **96.65** | **16.91** |

Performance was record dependent. In particular, the Record 106 window at 130 –140 s showed reduced R-peak sensitivity of 66.67%, which contributed to missed downstream PVC events. These results indicate that QRS-detection errors represent one source of error propagation into the PVC-classification stage.

RR intervals calculated from consecutive detections were subsequently converted to instantaneous heart rate. Fig. 5 shows representative stable sinus-rhythm behavior from Record 100; RR intervals were mainly approximately 0.77–0.87 s, corresponding to approximately 69–78 beats/min.

**Fig 5.**
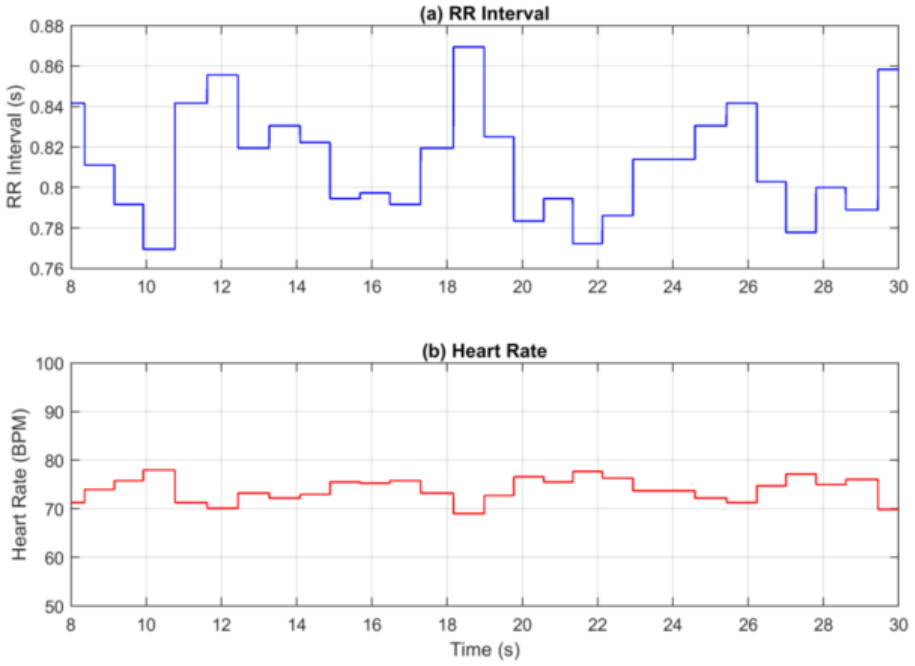
RR-interval and instantaneous heart-rate trends for MIT-BIH Record 100.

### 4.3. Rhythm classification

Representative tests showed normal rhythm in Record 100 at approximately 69.9 beats/min, bradycardic intervals in Record 101 including approximately 59.67 beats/min, and tachycardic operation in Record 213 including approximately 114.3 beats/min. Fig. 6 illustrates the three rhythm examples and their fixed decision boundaries, and Table 2 summarizes the validation.

**Table 2.** Representative rhythm-classification validation.

| **Rhythm** | **MIT-BIH segment** | **Representative HR** | **Output** |
|---|---|---|---|
| Normal | Record 100, 0–30 s | 69.9 BPM | Correct |
| Bradycardia | Record 101, 280–310 s | 59.67 BPM | Correct |
| Tachycardia | Record 213, 0–30 s | 114.3 BPM | Correct |

**Fig 6.**
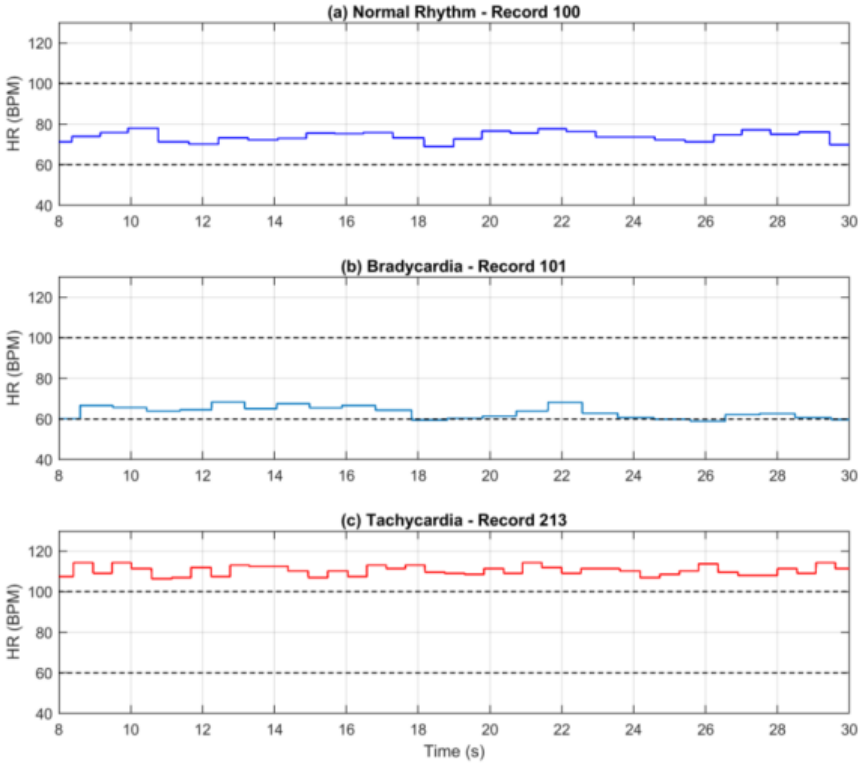
Heart-rate-based normal, bradycardic, and tachycardic rhythm examples.

### 4.4. PVC detection example

A representative PVC result for Record 119 is shown in Fig. 7. In the illustrated 20 –30 s segment, all five expert-annotated PVCs were detected without a false-positive event. This window belongs to the development set (Table 3) and is shown only to illustrate detector output; it is not used to characterize held-out performance.

**Table 3.** PVC detection on the three development windows (used for threshold selection; not included in the held-out estimates).

| **Record** | **Window (s)** | **Ref. PVCs** | **TP** | **FP** | **FN** | **Sens. (%)** | **Prec. (%)** | **F1 (%)** |
|---|---|---|---|---|---|---|---|---|
| 106 | 88–98 | 5 | 4 | 2 | 1 | 80.00 | 66.67 | 72.73 |
| 119 | 20–30 | 5 | 5 | 0 | 0 | 100.00 | 100.00 | 100.00 |
| 200 | 0–10 | 8 | 7 | 1 | 1 | 87.50 | 87.50 | 87.50 |
| **Total** | **3 windows** | **18** | **16** | **3** | **2** | **88.89** | **84.21** | **86.49** |

**Fig 7.**
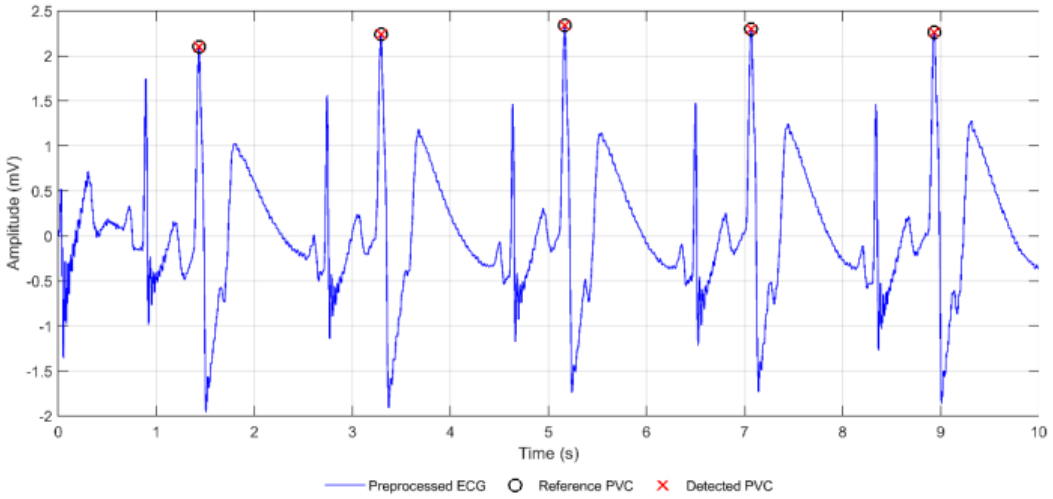
PVC detection example from MIT-BIH Record 119 (development window, 20 –30 s) with expert reference and detected PVC markers; time is relative to the window start.

### 4.5. Held-out PVC evaluation

The held-out PVC evaluation comprised 56 expert-annotated PVCs from nine non-overlapping 10-s windows obtained from MIT-BIH Records 106, 119, and 200 (Table 4). These windows were not used during development or threshold selection.

**Table 4.** Held-out PVC detection performance across nine non-overlapping 10-s windows.

| **Record** | **Window (s)** | **Ref. PVCs** | **TP** | **FP** | **FN** | **Sens. (%)** | **Prec. (%)** | **F1 (%)** |
|---|---|---|---|---|---|---|---|---|
| 119 | 190–200 | 6 | 5 | 0 | 1 | 83.33 | 100.00 | 90.91 |
| 119 | 290–300 | 5 | 5 | 1 | 0 | 100.00 | 83.33 | 90.91 |
| 119 | 460–470 | 5 | 4 | 2 | 1 | 80.00 | 66.67 | 72.73 |
| 119 | 760–770 | 5 | 4 | 2 | 1 | 80.00 | 66.67 | 72.73 |
| 119 | 960–970 | 5 | 4 | 2 | 1 | 80.00 | 66.67 | 72.73 |
| 119 | 1130–1140 | 5 | 5 | 1 | 0 | 100.00 | 83.33 | 90.91 |
| 200 | 1100–1110 | 9 | 4 | 0 | 5 | 44.44 | 100.00 | 61.54 |
| 200 | 70–80 | 8 | 6 | 1 | 2 | 75.00 | 85.71 | 80.00 |
| 106 | 130–140 | 8 | 3 | 0 | 5 | 37.50 | 100.00 | 54.55 |
| **Pooled** | **9 windows** | **56** | **40** | **9** | **16** | **71.43** | **81.63** | **76.19** |

The frozen classifier produced 40 true positives, 9 false positives, and 16 false negatives, corresponding to 71.43% sensitivity, 81.63% precision, and 76.19% F1 score. By record, sensitivity, precision, and F1 score were 37.50%, 100.00%, and 54.55% for Record 106 (8 PVCs); 87.10%, 77.14%, and 81.82% for Record 119 (31 PVCs); and 58.82%, 90.91%, and 71.43% for Record 200 (17 PVCs). The three development windows (Table 3) yielded 88.89% sensitivity, 84.21% precision, and 86.49% F1 score, but they were used for threshold selection and are not included in the held -out estimates.

The higher precision relative to sensitivity indicates that the detector was comparatively conservative: reported PVC candidates were often correct, but a meaningful fraction of expert-annotated PVCs were missed.

The most challenging windows occurred in Records 106 and 200. Quantitative R-peak analysis demonstrated that part of this loss arose upstream, because missed QRS complexes prevented subsequent RR- and QRS-energy-based PVC decisions. This interaction highlights the dependence of beat-level arrhythmia classification on robust QRS localization.

## 5. Discussion

The results demonstrate that a complete ECG-processing and arrhythmia-monitoring pipeline can be implemented using deterministic DSP operations and transparent rule-based logic within MATLAB/Simulink. The principal benefit is not maximum classification accuracy, but integration of preprocessing, QRS detection, RR analysis, rhythm classification, PVC feature extraction, reference validation, and dashboard visualization in a single observable environment.

A significant engineering finding emerged during preprocessing development. Minimum-order FIR filtering introduced approximately 4 s total delay, inconsistent with responsive monitoring. Replacing the FIR stages with IIR implementations substantially reduced observed delay while maintaining useful ECG morphology and spectral suppression. This demonstrates an implementation trade-off between linear-phase FIR filtering and lower-latency IIR processing in streaming biomedical applications.

The QRS detector serves as a common timing foundation for the downs treamrhythm and PVC-analysis stages and follows established low-complexity principles rather than claiming QRS-method novelty [1]. The heart-rate-based rhythm classifier remains fully traceable to measured RR intervals, although rate thresholds alone cannot distinguish the full range of clinically relevant rhythms.

PVC classification was substantially record dependent. The held-out evaluation achieved 71.43% sensitivity, 81.63% precision, and 76.19% F1 score, with lower sensitivity than on the development windows (88.89%), consistent with threshold selection on a small development set. Record 119 was strongest (81.82% F1), while Record 106 was most challenging (54.55% F1). Fixed RR- and energy-ratio thresholds therefore do not generalize equally across ventricular morphologies and rhythm patterns.

Quantitative evaluation of the upstream R-peak detector provides additional insight into the PVC results. Across the samenine held-out windows, R-peak detection achieved 94.39% sensitivity, 99.02% positive predictivity, and 96.65% F1 score, with a mean absolute timing error of approximately 16.91 ms. The reduced R-peak sensitivity observed in the challenging Record 106 window indicates that some PVC false negatives originated from missed QRS events rather than from PVC decision logic alone.

Previous RR-based PVC research supports the physiological usefulness of prematurity and compensatory-pause characteristics [5]. The QRS-energy ratio in the present work provides a lightweight morphology-related feature using signals already available in the QRS pathway. Richer Bayesian morphology tracking [6] or learned representations [7], [8] can exploit more ECG information but at greater algorithmic or training complexity.

The held-out precision of 81.63% exceeded the sensitivity of 71.43%, indicating a relatively conservative detector. The 16 false negatives are therefore the principal classification limitation, and future improvements should prioritize sensitivity without substantially increasing the 9 observed false positives.

## 6. Limitations

First, the final held-out PVC evaluation contained 56 PVCs from selected high-event-density segments of three MIT-BIH records, rather than a random or prevalence-weighted sample of the entire database. The performance estimates should therefore be interpreted as targeted stress-test results rather than whole-database generalization [2]–[4].

Second, the held-out windows were selected deterministically based on annotated PVC density to maximize the number of ventricular events within the practical simulation budget. Although this procedure increases the statistical usefulness of short simulations, it may bias the evaluation toward PVC-rich and potentially more abnormal intervals. The distribution across records was also imbalanced (8 PVCs from Record 106, 31 from Record 119, and 17 from Record 200), so pooled metrics are influenced by record-specific morphology.

Third, the three initial windows containing 18 PVCs were used during development and were excluded from the main held-out performance estimates.

PVC detection uses fixed RR-ratio and QRS-energy-ratio thresholds. These were frozen before held-out evaluation, avoiding record-specific retuning, but cannot account for all inter-patient differences in rate, morphology, noise, or ectopic-beat characteristics.

The rhythm classifier considers only normal rhythm, bradycardia, and tachycardia using instantaneous heart-rate boundaries. It does not diagnose atrial fibrillation, ventricular tachycardia, supraventricular ectopy, conduction disorders, or other complex rhythms.

The present study uses prerecorded ECG in Simulink simulation, and no claim of hardware real-time execution is made. Embedded execution time, memory, power, and end-to-end hardware latency were not measured, and analysis was primarily single-channel MLII.

The ±100 ms event-matching tolerance was applied consistently, but alternative matching protocols may yield different results; direct comparison with other publications requires consideration of dataset partitioning, annotation handling, and matching rules.

## 7. Conclusion and future work

This study presented an interpretable streaming-oriented ECG-processing architecture developed in MATLAB/Simulink for preprocessing, QRS/R-peak detection, RR-interval and heart-rate estimation, rhythm classification, and PVC detection. The final preprocessing architecture used low-latency IIR filtering after experimental evaluation demonstrated excessive delay from minimum-order FIR filtering.

The system demonstrated transparent classification of representative normal, bradycardic, and tachycardic rhythm conditions. PVC detection used two lightweight features: RR-interval ratio and QRS-energy ratio, combined through fixed rule-based logic.

After algorithm development on three initial windows containing 18 annotated PVCs, the frozen detector was evaluated on nine additional non-overlapping 10-s windows containing 56 expert-annotated PVCs. The held-out evaluation produced 40 true positives, 9 false positives, and 16 false negatives, corresponding to 71.43% sensitivity, 81.63% precision, and 76.19% F1 score. The upstream R-peak detector achieved 94.39% sensitivity, 99.02% positive predictivity, and 96.65% F1 score across the same held-out windows, with approximately 16.91 ms mean absolute timing error. Record-dependent variation demonstrates both feasibility and limitations of fixed-threshold low-complexity PVC classification.

Future work should investigate adaptive RR and morphology thresholds, signal-quality-aware decision logic, improved QRS morphology descriptors, larger patient-independent evaluation, multi-lead fusion, more complex rhythm detection, systematic noise-stress testing, fixed-point conversion, generated C/C++ code, and hardware/processor-in-the-loop validation.

## Data and code availability

The ECG recordings and expert annotations analyzed in this study are publicly available through the MIT-BIH Arrhythmia Database on PhysioNet [3], [4]. The MATLAB/Simulink model, evaluation scripts, and figure-generation scripts are available in a public repository; the link is provided in the title page file to preserve anonymity during review.

## Declarations CRediT authorship contribution statement

Abdullah Atif: Conceptualization, Methodology, Software, Investigation, Writing – original draft. Mst. Jannatul Ferdous: Supervision, Writing – review & editing.

## Funding

This research did not receive any specific grant from funding agencies in the public, commercial, or not-for-profit sectors.

## Declaration of competing interest

The authors declare that they have no known competing financial interests or personal relationships that could have appeared to influence the work reported in this paper.

## Ethics statement

This study used only publicly available, de-identified recordings from the MIT-BIH Arrhythmia Database; no new data were collected from human participants.

## Declaration of generative AI and AI-assisted technologies in the manuscript preparation process

During the preparation of this work the authors used Claude (Anthropic), ChatGPT (OpenAI), and Microsoft Copilot in order to improve language and readability, check consistency, and format the manuscript. After using these tools, the authors reviewed and edited the content as needed and take full responsibility for the content of the publication.

## Data availability

The ECG recordings and expert annotations analyzed in this study are publicly available through the MIT-BIH Arrhythmia Database on PhysioNet (https://doi.org/10.13026/C2F305).

## Code and model availability

The MATLAB/Simulink model, evaluation scripts, and figure-generation scripts are available at https://github.com/atif721/ECG_Simulink_Project and archived on Zenodo (https://doi.org/10.5281/zenodo.24280976). The ECG data are available from PhysioNet (see above).

## Notes

### Competing Interest Statement

The authors have declared no competing interest.

### Author Declarations

This study used only pre-existing, de-identified, publicly available human ECG data from the MIT-BIH Arrhythmia Database hosted by PhysioNet. The data and expert beat annotations were publicly available before initiation of this study. No participants were recruited, no new human data or biological samples were collected, and the authors had no access to direct participant identifiers. The dataset is available at: https://physionet.org/content/mitdb/1.0.0/

## References

[1] J. Pan, W.J. Tompkins, A real-time QRS detection algorithm, IEEE Trans. Biomed. Eng. BME-32 (3) (1985) 230–236, 10.1109/TBME.1985.325532.

[2] G.B. Moody, R.G. Mark, The impact of the MIT-BIH Arrhythmia Database, IEEE Eng. Med. Biol. Mag. 20 (3) (2001) 45–50, 10.1109/51.932724.

[3] [dataset] G. Moody, R. Mark, MIT-BIH Arrhythmia Database, version 1.0.0, PhysioNet, 2005, 10.13026/C2F305.

[4] A.L. Goldberger, L.A.N. Amaral, L. Glass, J.M. Hausdorff, P.C. Ivanov, R.G. Mark, J.E. Mietus, G.B. Moody, C.-K. Peng, H.E. Stanley, PhysioBank, PhysioToolkit, and PhysioNet: components of a new research resource for complex physiologic signals, Circulation 101 (23) (2000) e215–e220, 10.1161/01.CIR.101.23.e215.

[5] P. Cuesta, M.J. Lado, X.A. Vila, R. Alonso, Detection of prematureventricular contractionsusing the RR-interval signal: a simple algorithmfor mobile devices, Technol. Health Care 22 (4) (2014) 651–656, 10.3233/THC-140818.

[6] O. Sayadi, M.B. Shamsollahi, G.D. Clifford, Robust detection of premature ventricular contractions using a wave-based Bayesian framework, IEEE Trans. Biomed. Eng. 57 (2) (2010) 353–362, 10.1109/TBME.2009.2031243.

[7] J. Yu, X. Wang, X. Chen, J. Guo, Automatic premature ventricular contraction detection using deep metric learning and KNN, Biosensors 11 (3) (2021) 69, 10.3390/bios11030069.

[8] D. Kraft, G. Bieber, P. Jokisch, P. Rumm, End-to-end premature ventricular contraction detection using deep neural networks, Sensors 23 (20) (2023) 8573, 10.3390/s23208573.

